# Association of genetically-predicted lipid traits and lipid-modifying targets with the risk of heart failure

**DOI:** 10.1101/2022.03.24.22272860

**Authors:** Jun Xiao, Jianguang Ji, Xi Yang, Keyuan Chen, Liangwan Chen, Wuqing Huang

## Abstract

**Aims:** To assess the association of lipid traits and lipid-modification via licensed or investigational targets with heart failure (HF) risk using 2-sample Mendelian randomization (MR) study.

**Methods and results:** Genetic variants obtained from genome-wide association studies (GWASs) in UK Biobank as instrumental variables to investigate the association of lipid traits (low-density lipoprotein cholesterol (LDL-C), high-density lipoprotein cholesterol (HDL-C), triglyceride (TG), apolipoprotein B (ApoB) and apolipoprotein AI (ApoAI)) and lipid-modifying effect of eight licensed or investigational drug targets with HF risk by using the inverse-variance weighted method. In this study, we observed that genetically-predicted levels of LDL-C, TG, LDL-C and ApoB were significantly related to HF risk, which were mainly mediated by CHD. Further MR analyses identified PCSK9, CETP and LPL, but not HMGCR, as potential targets to prevent HF. The genetic proxy of LDL-C and ApoB increase modified by PCSK9 showed similar evidence in increasing risk of HF (P_LDL-C_=1.27*10^−4^; P_ApoB_=1.94*10^−4^); CETP played a role in HF risk via modifying all investigational lipid traits with the strongest evidence though ApoB (P=5.87*10^−6^); LPL exerted effects on HF via modifying most lipid traits with the strongest evidence observed via modifying TG (P=3.73*10^−12^).

**Conclusions:** This 2-sample MR study provided genetic evidence of the associations between lipid traits and HF risk, which were mostly mediated by CHD. Besides, drug target MR studies indicated that PCSK9 inhibition, CETP inhibition and LPL activation, but not statins, were effective in reducing HF risk.

## Introduction

Heart failure (HF) is a syndrome characterized by high morbidity and mortality (1). With an aging population, the incidence is rapidly growing globally and the lifetime risk is estimated to be around 20% (1). Although major advances have been made for its treatment, the prognosis of HF is still disappointing with a 5-year survival rate of around 50% (1). Primary prevention should be prioritized to reduce the public health burden from HF. A number of factors may contribute to the aetiology of HF, most notably CHD, hypertension, obesity and diabetes, indicating a variety of underlying pathogenesis (1). Dyslipidemia is frequently co-occur with these diseases as well as with HF, suggesting that dyslipidemia plays a role in HF and lipid-modifying therapies may provide a preventive strategy for HF (2, 3). Although dyslipidemia is an established risk factor of many cardiovascular diseases, especially CHD, findings from prior studies are mixed in terms of the association between lipid traits and HF (2, 3). Stains are a class of widely-used lipid-modifying drugs with Class I recommendation for primary and secondary prevention of atherosclerotic cardiovascular events; however, the role of statins in HF is still debatable (4, 5). Besides, a series of novel lipid-modifying agents are emerging, some of which have shown a desirable efficacy in reducing the risk of CHD, such as PCSK9 inhibitors, but the potential effect of these agents on HF is unclear.

Thus in this study, we performed two-sample Mendelian randomization (MR) analyses to explore the association of genetically-predicted lipid traits and lipid-modifying targets with HF risk. We first conducted univariable MR analyses to estimate the association between genetically-predicted lipid traits (circulating lipids and apolipoproteins) and the risk of HF. Next, multivariable MR analyses were further performed to assess the mediated role of common HF-related risk factors (CHD, type 2 diabetes, BMI, and blood pressure) in the observed relationships. At last, we performed drug target MR studies to explore the potential effective drug targets against HF by investigating the association between the genetically-predicted lipid-modifying effect of several well-known or novel targets and HF risk.

## Methods

Two sample MR analyses were conducted based on publicly available summary statistic data from genome-wide association studies (GWASs) (**Supplementary Table 1**). All these studies had approval from related institutional review boards and all participants in these studies provided informed consent.

## Data source

Summary-level GWAS data for lipid traits were obtained from the UK Biobank, which covered the largest-scale GWAS genotyping (number of SNPs=12,321,875) and lipid traits phenotyping, including low-density lipoprotein cholesterol (LDL-C, N=440,546), triglyceride(TG, N=403,943), high-density lipoprotein cholesterol (HDL-C, N=441,016), apolipoprotein B (ApoB, N=439,214) and apolipoprotein AI (ApoAI, N=393,193) (6). Analyses were performed in UK Biobank participants of European descent and included age, sex, and the type of genotyping chip (the UK BiLEVE array or the UKBB Axiom array) as covariates in regression models. Estimates from GWASs for these lipid traits are equivalent to a one standard deviation increase. Summary-level data for HF were obtained from the meta-analysis of HF GWASs, including 977,323 participants of European ancestry from 26 studies in the HERMES Consortium (cases=47,309 and controls=930,014). Participants with a clinical diagnosis of HF were defined as cases and participants without HF were defined as controls, and details of case definition for each study had been described previously (7). All analyses included in the meta-analysis have adjusted for age and sex, and meta-analysis was performed by using the inverse variance weighted (IVW) fixed-effect model. The details of GWAS summary data for covariates in multivariable MR analyses are presented in **Supplementary Table 1**.

### Selection of genetic instruments

To proxy the levels of lipid traits, we identified all SNPs associated with LDL-C, TG, HDL-C, ApoB or ApoAI at a genome-wide significance level (p<5.0□×□10^−8^) as instrumental variables (IVs), irrespective of the genomic position of SNPs. To ensure the independence of IVs, the identified genetic variants were filtered for linkage disequilibrium (LD) in both univariable and multivariable MR analyses as measured by r^2^ (r^2^□ <□0.001 within 10,000 kb window by using European reference panel from 1000 Genomes Project). All SNPs were harmonized via harmonization function in “TwoSampleMR” R package with default parameters. The details of IVs for LDL-C, TG, HDL-C, ApoB or ApoAI were presented in **Supplementary Tables 3 and 4**.

As shown in **Supplementary Table 2**, we further assessed the association of lipid modulation via licensed (HMGCR, PCSK9, NPC1L1 and PPARA) or investigational (CETP, LPL, ANGPTL3 and APOC3) drug targets with HF risk, including three drugs primarily for decreasing LDL-C level (HMGCR, PCSK9 and NPC1L1), four targets for decreasing TG level (PPARA, LPL, ANGPTL3, and APOC3), and one target for increasing HDL-C level (CETP). To predict the lipid-modifying effect of these drug targets, we selected SNPs within 100 kb windows from the region of each drug target gene that was associated with related lipid traits at a genome-wide significance level (p<5.0□×□10^−8^). GWAS summary data of lipid traits were also obtained from UK Biobank as mentioned above to identify these SNPs. Genetic instrument selection for these drug targets was based on summary data from the GWAS of LDL-C (to instrument HMGCR, PCSK9 and NPC1L1), TG (to instrument PPARA, LPL, ANGPTL3, and APOC3) and HDL-C (to instrument CETP). Given that ApoB is the major transporter for LDL-C/TG and ApoAI is the major transporter for HDL-C, we further generated genetic instruments for these drugs targets based on summary data from the GWAS of ApoB (to instrument HMGCR, PCSK9, NPC1L1, PPARA, LPL, ANGPTL3 and APOC3) and ApoAI (to instrument CETP). To maximize the instrument strength for each drug target, SNPs selected as instruments were allowed to be weak linkage disequilibrium (r^2^ <0.30 within 100 kb window by using European reference panel from 1000 Genomes Project) with each other.

If lipid-modifying effects via some drug targets were identified to be significantly associated with HF risk, we further generated genetic instruments for these drugs targets based on summary data from the GWAS of all lipid traits (LDL-C, TG, HDL-C, ApoB and ApoAI) to investigate if these drugs exerted their protective effect on HF via multiple lipid-modifying pathways. Next, to reveal how these targets affect lipid traits, we further investigated the association of genetically-predicted expression of these drug targets with each lipid trait level, which was estimated by combining eQTLs of drug targets and GWAS summary data of lipid traits. Furthermore, we also tested if there were associations between genetically-predicted expression of the identified drug targets and heart failure risk to provide additional evidence. The eQTLs summary-level data were retrieved from eQTLGen Consortium or GTEx Consortium V8, and estimates are equivalent to a one standard deviation increase. Only cis-eQTLs significantly (p<5.0□×□10^−8^) associated with the expression of drug target genes were selected as IVs, and eQTLs within 1□Mb on either side of the encoded gene were defined as cis-eQTLs.

### Statistical analyses

#### Primary analysis

TwoSampleMR R package was used to perform MR analyses, and IVW-MR method was applied to combine effect estimates. The univariable MR analyses were first performed to estimate the associations between genetically-predicted lipid traits and HF risk. Multivariable MR analysis was further performed to assess the potential mediated pathways between lipid traits and HF. CHD, type 2 diabetes, BMI, systolic and diastolic blood pressure are established risk factors of HF, thus we included these factors one by one in each multivariable MR model to investigate their potential mediation effect. Next, we conducted univariable and multivariable MR analyses to explore the genetically-predicted lipid-modifying effects mediated by each drug target on the risk of HF, in which estimates were also derived by using the IVW-MR method. To account for multiple testing, Bonferroni correction was used to adjust the thresholds of the significance level. All analyses were conducted in R software, version 4.1.0.

#### Sensitivity analysis

The F-statistic value was used to assess the strength of each IV (8). MR-PRESSO analysis was applied to test the horizontal pleiotropy of IVs, where P<0.05 in MR-PRESSO Global test indicates the presence of horizontal pleiotropy (9). MR-PRESSO analysis can further identify horizontal pleiotropic outliers and compare the estimates before and after outlier removal by using the distortion test.

Summary-data-based MR (SMR) method was used to estimate the association between the expression of drug targets and lipid traits using summary data from eQTL and GWAS studies (10). HEIDI test was applied to test the role of LD in the observed association, where P<0.01 indicates that association might be due to linkage (11). Analyses were conducted in SMR software, version 1.03.

## Results

### Genetic instruments selection and GWAS of HF

As shown in **Supplementary File 1**, the number of SNPs ranged from 47 to 311 to instrument lipid traits, ranged from 1 to 90 to instrument lipid-modifying therapies. F-statistics for all genetic instrument variants were over 27, indicating minimized weak instrument bias in our study. GWAS of HF was derived from 26 studies (17 cohort studies and 9 case-control studies) from HERMES Consortium, including 47,309 cases and 930,014 controls.

### Association between genetically-predicted lipid traits and HF risk

As shown in **Figure 1 and Supplementary Table 3**, results from univariable MR analysis showed significant associations between most genetically-predicted lipid traits and the risk of HF (p<0.01), except for ApoAI with weak evidence (P=1.25*10^−02^). Higher level of genetically-predicted LDL-C (OR=1.16, P=3.58*10^−03^), TG (OR=1.14, P=2.63*10^−05^) and ApoB (OR=1.18, P=1.82*10^−05^) was associated with a higher incidence of HF, whereas higher level of genetically-predicted HDL-C (OR=0.89, P=3.45*10^−04^) and ApoAI (OR=0.93, P=1.25*10^−02^) was related to a lower risk of HF. However, in the multivariable MR analysis (**Figure 1 and Supplementary Table 4**), all these associations attenuated to null after being adjusted for CHD (P>0.05), suggesting a complete mediated effect of CHD in the association between lipid traits and HF risk. Most associations remained similar when adjusted for type 2 diabetes, BMI, or blood pressure. As shown in **Supplementary Table 3**, MR-PRESSO analysis suggested the presence of horizontal pleiotropy for all reported results (P<0.05) while distortion test showed no difference in the estimates before and after outlier removal (P>0.05).

**Figure 1.**
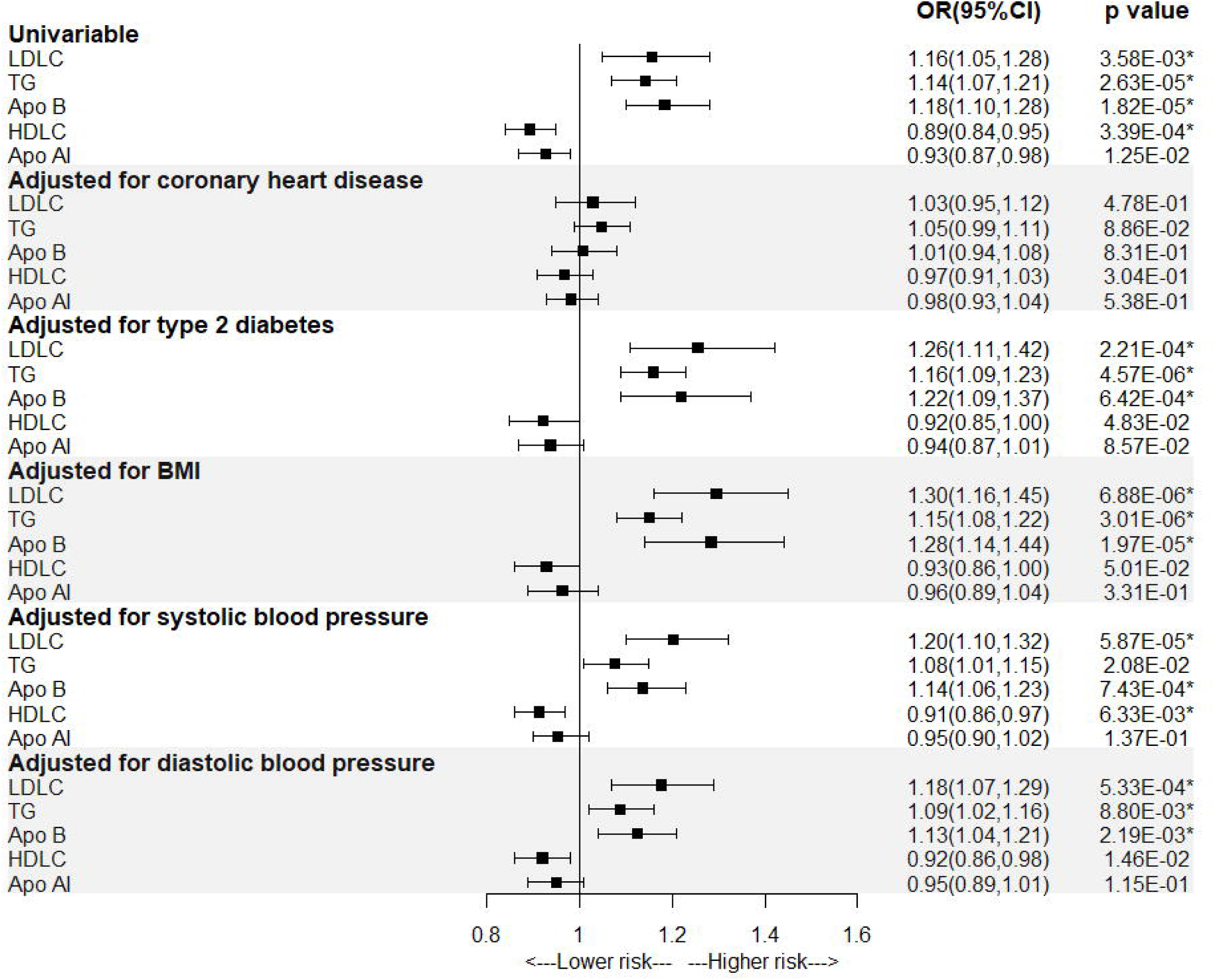
Association between genetically-predicted lipid traits and the risk of heart failure. *p<0.01 indicates significance after bonferreni correction.

### Association between the genetically-predicted primary lipid-modifying effect of targets and HF risk

The detailed information of lipid-modifying drug targets is presented in **Supplementary Table 2**. As shown in **Figure 2 and Supplementary Table 5**, three lipid-modifying targets were shown to have genetically-predicted effects on the risk of HF, including PCSK9, CETP and LPL, while no association was found of genetically-predicted LDL-C level modified by HMGCR (the target gene of statins) with HF risk (OR=0.88, P=0.121). Significant positive associations with HF risk were found for genetically-predicted LDL-C modified by PCSK9 (OR=1.28, P=1.27*10^−4^), genetically-predicted TG modified by LPL (OR=1.22, P=3.73*10^−12^), and genetically-predicted ApoB modified by PCSK9 and LPL (OR_PCSK9_=1.27, P=1.94*10^−4^; OR_LPL_=1.98, P=5.54*10^−7^). Higher genetically-predicted HDL-C and ApoAI modified by CETP was related to a lower risk of HF (OR_HDL-C_ =0.93, P=1.17*10^−5^; OR_ApoAI_ =0.92, P=4.85*10^−5^).

**Figure 2.**
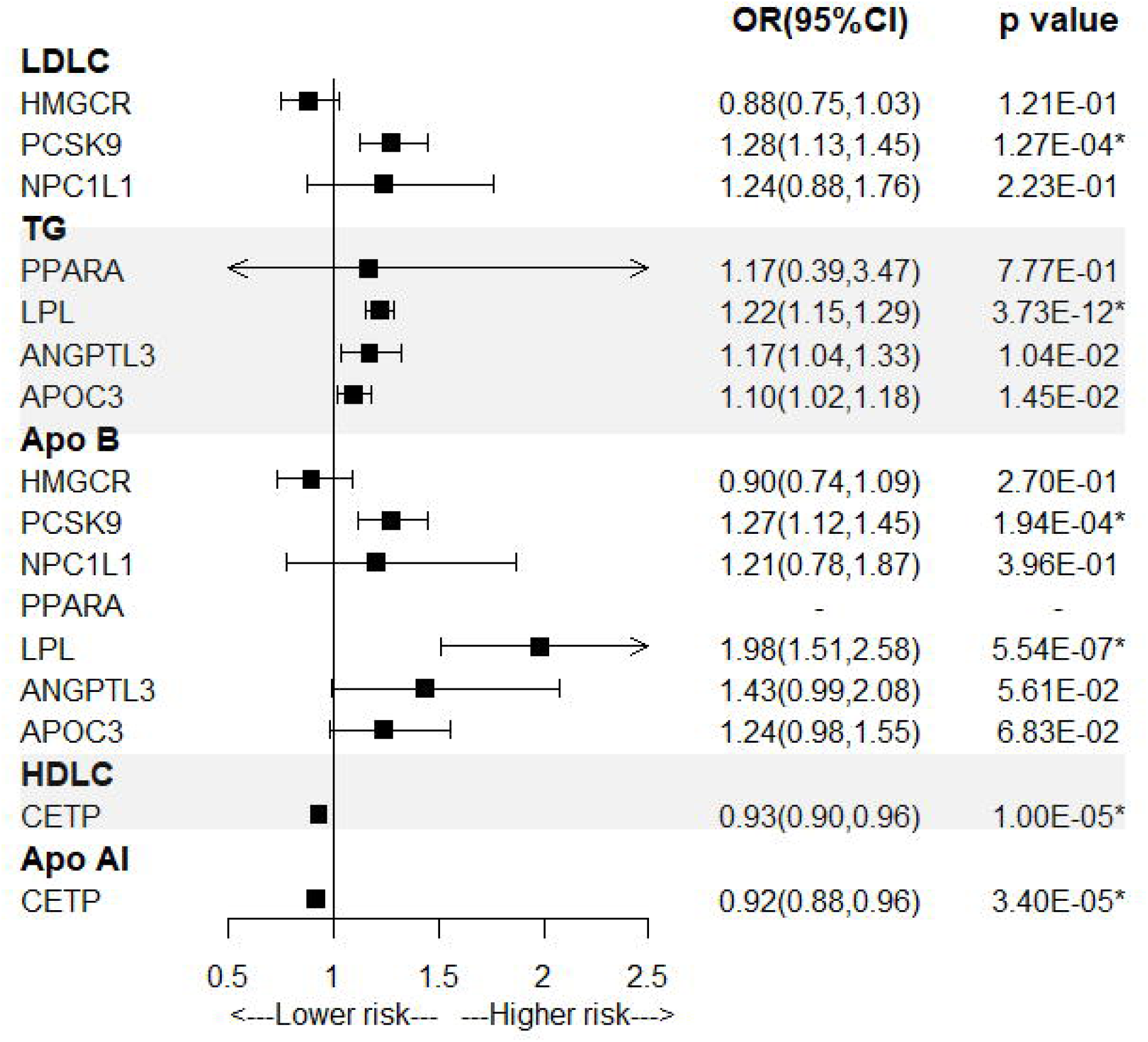
Association between genetically-predicted primary lipid-modifying effect of targets and the risk of heart failure. *p<0.00625 indicates significance after bonferreni correction.

### Association between genetically-predicted lipid-modifying effects of identified targets and HF risk

We further investigated if PCSK9, CETP or LPL played a role in HF via modifying multiple lipid traits. Available instruments variants were identified mainly in LDL-C and ApoB GWAS to instrument PCSK9, and a significant positive association was found between LDL-C/ApoB level modified by PCSK9 and risk of HF (**Figure 3 and Supplementary Table 6**); similarly, SMR analyses showed that the higher expression of PCSK9 gene was significantly associated with the higher level of LDL-C (beta=0.227236, P=3.24*10^−12^) and ApoB (beta=0.217571, P=4.18*10^−12^), and a weaker evidence was observed between higher PCSK9 expression and higher HDL-C level (P=2.37*10^−03^) (**Figure 4 and Supplementary Table 8**); these findings suggested that PCSK9 inhibition may reduce HF risk via decreasing LDL-C and ApoB level. Available instrument variants were identified across all these lipid traits GWASs to instrument CETP, and a higher level of LDL-C/TG/ApoB and lower level of HDL-C/ApoAI modified by CETP were associated with a higher risk of HF; SMR analyses showed that the higher expression of CETP gene was significantly associated with the higher level of LDL-C (P=2.89*10^−24^), TG (P=1.03*10^−26^) and ApoB (P=3.68*10^−35^), and with the lower level of HDL-C (P=3.02*10^−57^) and ApoAI (P=2.67*10^−56^); these results revealed that CETP inhibition may play a protective effect on HF via modifying all these lipids traits (decreasing LDL-C/TG/ApoB and increasing HDL-C/ApoAI). Available instrument variants were identified across most lipid traits GWASs except in LDL-C GWAS to instrument LPL, and a higher level of TG/ApoB and lower level of HDL-C/ApoAI modified by LPL were associated with a higher risk of HF; SMR analyses showed that the higher expression of LPL gene was significantly associated with the lower level of TG (P=4.14*10^−22^) and ApoB (P=3.37*10^−11^), with the higher level of HDL-C (P=5.36*10^−22^) and ApoAI (P=3.14*10^−21^), and a weaker evidence with lower level of LDL-C (P=3.85*10^−3^); these results indicated that LPL activation may decrease HF risk via decreasing TG/ ApoB, and increasing HDL-C/ApoAI. However, all observed associations between genetically-predicted lipid-modifying effect and HF risk became weaker after being adjusted for CHD, in which the association of CETP-modified ApoB/ApoAI/HDL-C level and LPL-modified TG/HDL-C with HF risk remained significant (**Supplementary Table 7**). Results from SMR analysis for the association between PCSK9, CETP, LPL expression and HF provided additional evidence although some of which lacked statistical significance, in which relatively small sample size of eQTLs data may lead to insufficient statistical power (**Supplementary Table 1**). Consistent with results from drug target MR analyses above, SMR analyses showed that higher PCSK9/CETP expression was related to higher HF risk while higher LPL expression associated with lower HF risk, suggesting PCSK9 inhibition, CETP inhibition or LPL activation had a protective effect against HF (**Supplementary Table 9**).

**Figure 3.**
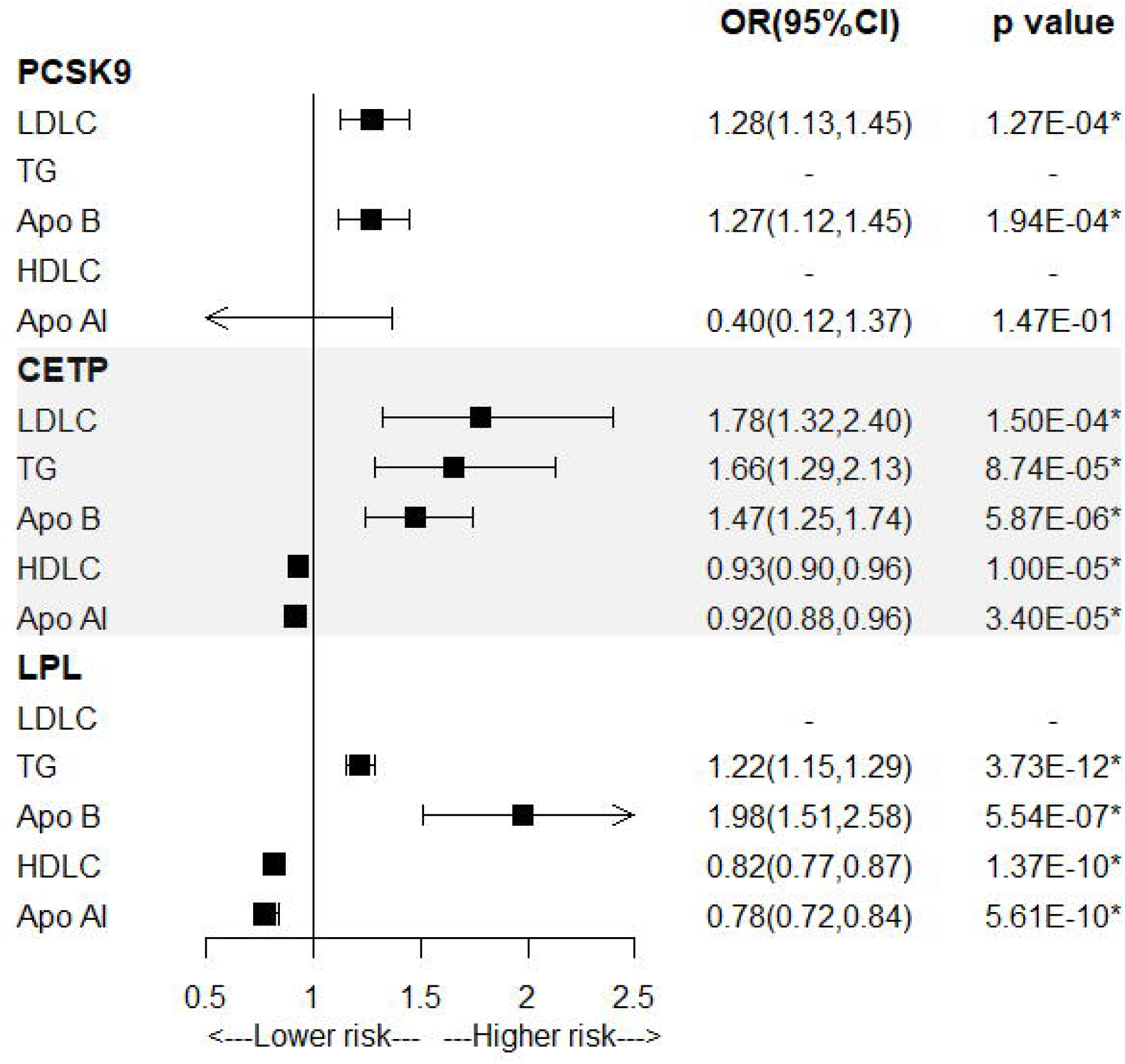
Association between genetically-predicted lipid-modifying effects of identified targets and the risk of heart failure. *p<0.00167 indicates significance after bonferreni correction.

**Figure 4.**
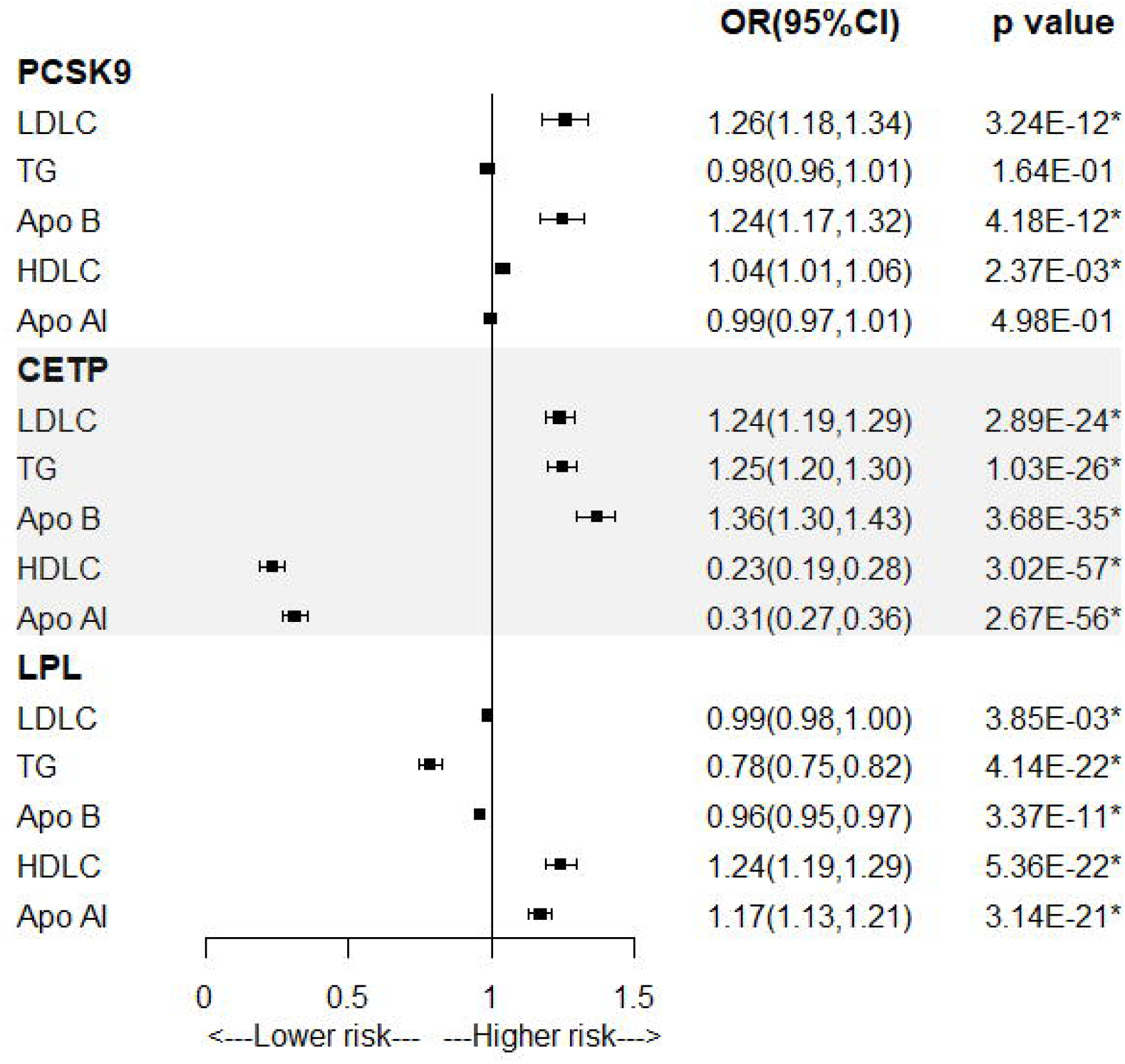
Association between genetically-predicted expression in whole blood of identified targets and the concentration of lipid traits. *p<0.00167 indicates significance after bonferreni correction.

## Discussion

In this study, we observed that genetically-predicted levels of lipid-traits (LDL-C, TG, LDL-C, ApoB) were related to HF risk, which appeared to be mostly mediated by CHD while type 2 diabetes, BMI and blood pressure played a small role in the observed associations. Next, we further explored the genetically-predicted effects of modifying lipid traits via several drug targets that are licensed or under investigation. Three drug targets were identified significantly associated with HF risk, including PCSK9, CETP and LPL. Further analyses showed that PCSK9 inhibition reduced the HF risk mainly through decreasing the concentration of LDL-C and ApoB, CETP inhibition played a protective effect on HF via modifying all investigational lipid traits and LPL activation via modifying most lipid traits except LDL-C.

The association between lipid traits and HF were relatively limited with mixed resultsA study of 40,607 patients with myocardial infarction (MI) found that LDL-C reduction significantly decreased the incidence of HF hospitalization (12), while a recent study including 103,860 individuals did not observe an association between LDL-C concentration and the HF risk (13). A community-based prospective study found that lower HDL-C and a higher ratio of apolipoprotein B/A-1 were independent risk factors of HF risk (14). Results from the Framingham Heart Study showed that dyslipidemia was associated with HF risk independent of MI, in which an elevated non-HDL-C or a lower HDL-C at baseline was related to a higher risk of HF (15). While another study from the Multiethnic Study of Atherosclerosis found a mediated role of myocardial infarction between lipids and incident HF (16). It is well-proven that omega-3 fatty acids can lower TG levels, and emerging evidence suggested that omega-3 fatty acid supplementation had benefits in reducing HF risk, indicating a negative association between TG level and HF risk(17). In the present MR study, we reported the association of genetically-predicted lipid traits and HF risk by combining the latest and largest-scale GWASs data. We found that there were significant associations between most genetically-predicted lipid traits level and incident HF (i.e., increasing LDL-C, TG or ApoB and decreasing HDL-C associated with an elevated incidence of HF), except for a weak negative association with genetically-predicted ApoAI concentration. It is noteworthy that these associations attenuated to null after being adjusted by CHD but remained similar after adjusted by diabetes, BMI or blood pressure, indicating that the relations between lipid traits and HF risk might be mostly mediated by CHD and lipid-modifying treatment mainly contributed to reducing the risk of atherosclerosis-related HF.

HMG-CoA reductase inhibitors (known as statins, encoded by HMGCR) are widely-used lipid-lowering drugs globally for coronary artery diseases prevention and treatment primarily through lowering LDL-C concentration. However, the role of statins in HF remains a subject of debate (4, 5). Two previous large-scale trials did not observe any benefits from statins treatment in the clinical outcomes of HF patients (18, 19). While a latest collaborative meta-analysis of up to 17 primary- and secondary-prevention trials revealed that statins modestly decreased the hospitalization rate due to non-fatal HF and the composite of HF outcomes but not HF mortality, and the risk reduction was independent of MI (20). The present study did not provide evidence for the benefits of statins in incident HF reduction, in which no significant association was found between genetically-predicted HMGCR-modified LDL-C and HF risk. Nevertheless, another three lipid-modifying targets were identified in this study that were significantly related to HF risk, including PCSK9, CETP and LPL.

Proprotein convertase subtilisin/kexin type 9 (PCSK9) protein is known to reduce LDL-C clearance via controlling low-density lipoprotein receptor (LDLR) recycling. A number of studies have demonstrated the desirable benefits of PCSK9 inhibitors in treating atherosclerosis diseases, but the effect on HF is uncertain. A multicenter prospective study observed that increasing level of PCSK9 protein was associated with an elevated risk of mortality (HR=1.24, 95%CI=1.04-1.49) and the composite outcomes (HR=1.21, 95%CI=1.05-1.40) in HF patients (21); prior MR study provided genetic evidence for the protective effect of lower PCSK9 protein concentration on HF risk (OR=0.79, 95% CI=0.71-0.87) (22); both suggesting the potential benefits of PCSK9 inhibition against HF. However, a meta-analysis with published trials did not observe a risk reduction of HF in patients who received PCSK9 inhibitors, while the insufficient follow-up in clinical trials may preclude from discovering the underlying effect (23). Findings from the current study indicated that PCSK9 inhibition contributed to reducing the risk of HF via decreasing the level of LDL-C and ApoB which were dependent on CHD, as there were significant positive associations between genetically-predicted LDL-C and ApoB level modified by PCSK9 and HF risk, but these associations attenuated to null after adjusted by CHD.

In recent years, cholesteryl ester transfer protein (CETP) inhibitors were developed for cardiovascular diseases treatment mainly via increasing HDL-C. However, results from current clinical trials are disappointing in terms of the therapeutic efficacy for cardiovascular diseases though being effective in modifying multiple lipid traits, none of CETP inhibitors have been approved. In contrast, MR studies generated consistent evidence regarding the benefits of reducing cardiovascular events by inhibiting CETP (22, 24, 25). Schmidt AF, et al. compared evidence derived from clinical trials and drug target MR study of CETP protein, showing that failures of CETP inhibitors in trials might be related to suboptimal target inhibition, off-target effects, or insufficiently follow-up, but not the target itself (22). On-target CETP inhibition, assessed by MR study, was expected to reduce the risk of adverse cardiovascular events, including HF (OR=0.96, 95% CI=0.93–0.98) (22). Brian A, et al. performed MR analysis based on 14 studies with 102837 participants, revealing that CETP inhibition led to a concordant decrease in LDL-C and ApoB concentration independent with HMGCR inhibition and the clinical benefit of decreasing LDL-C levels for cardiovascular diseases reduction probably depended on the absolute reduction in lipoprotein particles containing ApoB (25). Results from our study also suggested that CETP inhibition was effective for reducing incident HF via modifying multiple lipid traits. When adjusted for CHD, all observed associations became weaker while associations between genetically-predicted CETP-modified ApoB/HDL-C/ApoAI concentration and HF risk kept significant, indicating other underlying pathways apart from through atherosclerosis.

Lipoprotein lipase (LPL) is an enzyme hydrolyzing TG in triglyceride-rich lipoproteins (26). LPL activation contributes to lower TG levels and LPL deficiency would lead to hypertriglyceridemia (26). LPL is widely distributed in the heart and considered as the only enzyme to generate lipid-derived fatty acids that provide nearly 70% cardiac energy, thus LPL has a central role in cardiac energy homeostasis (27). In vivo experiments found LPL expression was downregulated in HF mice and LPL knockout hearts were susceptible to HF (27, 28). LPL has been considered as a novel potential target for dyslipidemia treatment and there are some investigational agents targeting LPL activation (29). Previous MR analyses showed that genetic proxied enhancing LPL activity were related to a reduced risk of CHD, but lacking evidence about the role in HF (30, 31). In this study, we found that genetically-predicted lipid traits change modified by LPL activation was also associated with lower risk of HF, including decreasing TG/ApoB and increasing HDL-C/ApoAI, most of which were partly mediated by CHD except for while associations with LPL-modified TG/HDL-C remained significance.

The use of genetic variants to instrument lipid traits and lipid-modifying targets contributed to minimizing bias from confounding and avoiding reverse causation. The latest GWASs of lipid traits with a coverage of the large-scale SNPs allowed us to generate genetic instruments for these drug targets. We additionally considered common risk factors of HF to identify possible mediating pathways between lipids and HF by performing multivariable MR analyses, and identified CHD as an important mediator. SMR analyses combining drug targets eQTLs and HF GWAS data could provide additional evidence for the potential effect of the identified drug targets on lipid traits and HF.

There are several limitations in this study. Firstly, this study explored the genetically-predicted lipid-modifying effect of drug targets on HF risk, which may not be equivalent to lipid-modifying therapies due to a gap between targets and available agents. However, these findings could provide some clues for HF prevention, and further clinical studies are needed to confirm these findings. Secondly, eQTLs data are limited, we performed SMR analyses combining available drug targets eQTLs and HF GWAS data, which generated inconsistent results between tissues. For example, genetically-predicted higher CETP expression in adipose tissue and small intestine but not in blood and heart were significantly related to a higher HF risk; genetically-predicted lower LPL expression in adipose tissue but not in blood was significantly associated with a higher HF risk; no significant association was found between genetically-predicted PCSK9 expression in blood or adipose tissue and HF risk. However, all these associations showed expected similar trends despite some without statistical significance, in which insufficient statistical power cannot be ruled out. Thirdly, summary-level GWAS data used in this study were mainly obtained from a population of European ancestry resulting in limited generalization.

## Conclusions

In conclusion, this MR study suggested a relationship between the genetically-predicted level of lipid traits (LDL-C, TG, HDL-C and ApoB) with HF risk, and three lipid-modifying drug targets were identified that might be effective to reduce HF incidence, including PCSK9 inhibition, CETP inhibition and LPL activation. These observed associations were mostly mediated by CHD, indicating that lipid-modifying treatments were more likely to have benefits in preventing CHD-related HF. Further studies are needed to test the observed relationships.

## Data Availability

All data produced in the present study are available upon reasonable request to the authors.

## Abbreviations

ANGPTL3: Angiopoietin Like 3
APOC3: Apolipoprotein C3
CETP: Cholesterylester Transfer Protein
CI: confidence interval
eQTLs: expression quantitative trait loci
GWAS: genome-wide association study
HF: heart failure
HEIDI: heterogeneity in dependent instruments
HMGCR: HMG-CoA reductase
IVW-MR: inverse-variance weighted Mendelian Randomization
LDL: low-density lipoprotein
LPL: Lipoprotein Lipase
MR-PRESSO: Mendelian Randomization Pleiotropy RESidual Sum and Outlier
NPC1L1: Niemann-Pick C1-Like 1
OR: odds ratio
PCSK9: Proprotein Convertase Subtilisin/Kexin type 9
PPARA: Peroxisome Proliferator Activated Receptor Alpha
SNP: single-nucleotide polymorphism
SMR: summary-data-based Mendelian Randomization.

## Acknowledgments

We thank the patients and investigators who contributed to the UK Biobank, HERMES Consortium, CARDIoGRAMplusC4D, DIAGRAM Consortium, GIANT Consortium, International Consortium of Blood Pressure, eQTLGen Consortium, and GTEx Consortium.

